# Glucagon-like Peptide 1 Receptor Agonists and Risk of Pulmonary Hypertension

**DOI:** 10.1101/2025.09.18.25336100

**Authors:** Jonah D. Garry, Suman Kundu, Chuck Alcorn, Svetlana Eden, Emily Smith, Robert Greevy, Hilary A. Tindle, Matthew S. Freiberg, Evan L. Brittain

## Abstract

**Background:** Cardiometabolic disease is a leading cause of pulmonary hypertension (PH), a progressive condition that increases the risk of death and heart failure hospitalization. Preventive therapies are lacking, and Glucagon-like peptide 1 receptor agonists (GLP-1 RAs) are a promising intervention due to their broad cardiometabolic benefits. Therefore, this study was designed to determine whether GLP-1 RA exposure is associated with reduced risk for developing PH.

**Methods:** We conducted a retrospective cohort study using data from the Veterans Health Administration. Patients with diabetes and an echocardiogram without PH who initiated a GLP-1 RA or Dipeptidyl Peptidase 4 inhibitor (DPP-4i) between January 1, 2007 and December 31, 2021 were included in the study. We used the active comparator, new user design to emulate a pragmatic clinical trial and employed inverse probability weighting (IPW) to adjust for confounding. GLP-1 RA new users were compared to DPP-4i new users. Patients were followed from first prescription date (baseline) until PH diagnosis, death, or medication cessation (latest date April 30, 2022). The primary outcome was development of PH, defined by an echocardiographic estimated right ventricular systolic pressure of 35 mmHg or greater.

**Results:** We identified 4,109 GLP-1 RA users and 7,384 DPP-4i users without PH at baseline. In comparison to DPP4i users, GLP-1 RA users were younger (median [Q1-Q3], 69 [63-73] vs 70 [65-74]) and similar in sex distribution (94.6% vs 95.6% male). Unadjusted incidence rates of PH for GLP-1 RA compared to DPP-4i were 54.8 vs 69.7 cases per 1000 person-years. IPW achieved covariate balance between groups (all standardized mean differences <0.10). In Cox regression models on the IPW cohort, GLP-1 RA exposure compared to DPP-4i exposure was associated with a 28% reduced risk of developing PH (HR, 0.72; 95%CI 0.61–0.84). Reduced risk of PH associated with GLP-1 RA use was consistent across sensitivity and subgroup analyses, including when stratified by obesity and diabetes severity.

**Conclusions:** GLP-1 RA use was associated with a reduced risk of developing PH. This finding supports the need for clinical trials and mechanistic studies to evaluate the potential role of GLP-1 RA therapy in the prevention of PH.

## Introduction

Pulmonary hypertension (PH) is a highly prevalent condition that most often arises from cardiopulmonary disorders.^1^ Regardless of etiology, both prevalent and incident PH are strongly associated with increased mortality risk.^2–4^ Despite the substantial population-level burden of PH and the large number of patients with cardiopulmonary disease at risk for its development there remain no clinical interventions proven to reduce the risk of PH.

Growing evidence implicates cardiometabolic risk factors as major contributors to the development and progression of PH in the general population.^2^ In this context, glucagon-like peptide 1 receptor agonists (GLP-1 RAs) have gained attention not only for their established benefits in glycemic control, weight management, and cardiovascular risk reduction but also as potential agents for treating and preventing PH.^5^ GLP-1 RAs may indirectly reduce pulmonary pressures by improving metabolic dysfunction and reducing passive pulmonary venous congestion from left sided heart failure. They may also exert direct protective effects on the pulmonary vasculature by reducing inflammation, enhancing nitric oxide signaling, and inhibiting endothelial-to-mesenchymal transition, pivotal processes implicated in PH pathogenesis.^6–10^

Given the broad therapeutic potential of GLP-1 RAs across various etiologies of PH, we designed this study to assess the association between GLP-1 RA exposure and hazard of developing PH in a large, nationwide cohort of US veterans with diabetes who were referred for echocardiography. We hypothesized that GLP-1 RA exposure would be associated with a lower risk of incident PH compared with Dipeptidyl Peptidase 4 inhibitor (DPP-4i) exposure.

## Methods

### Data Sources

This study was approved by the institutional review boards of Yale University, Veterans Affairs (VA) Connecticut Healthcare system, the Tennessee Valley Veterans Affairs (VA) Health Care System, and Vanderbilt University Medical Center. It has been granted a waiver of informed consent and is compliant with the Health Insurance Portability and Accountability Act.

This study utilized data collected as part of the Veterans Aging Cohort Study National (VACS-National) Cohort. Data was obtained from the United States Veterans Health Administration (VA) Corporate Data Warehouse and the VA Pharmacy Benefits Management database, which include information on over 13 million veterans. These resources include information on vital signs, health factors (e.g smoking data), diagnostic codes (International Classification of Diseases [ICD]), laboratory testing, documentation (including echocardiography [echo] reports) and prescription medications dispensed by VA pharmacies. Non-VA event data was obtained from Medicare and VA fee-for-service (Vista Fee Basis Package and the Program Integrity Tool) administrative ICD codes. Echo variables were abstracted from echo reports using a validated natural language processing tool that has been previously described.^11^ Data for this study was collected beginning on January 1, 2007, the year in which routine DPP-4i prescription began at the VA. The recruitment end date was 12/31/2021, after which data on medications prescribed through Medicare Part D is not available. Outcome data (echo reports) were available through April 30, 2022.

### Study Design

This was a retrospective cohort study designed to emulate a pragmatic clinical trial using an active comparator, new-user design.^12^ A visual depiction of the key study elements are displayed in Figure 1. New GLP-1 RA users were compared to new DPP-4i users -- the active comparator. DPP-4is are used to treat diabetes and act through a different mechanism than GLP-1 RAs. While there is no clinical evidence that DPP-4i affect pulmonary pressure, preclinical studies suggest that any potential effect is likely beneficial.^13^

**Figure 1:**
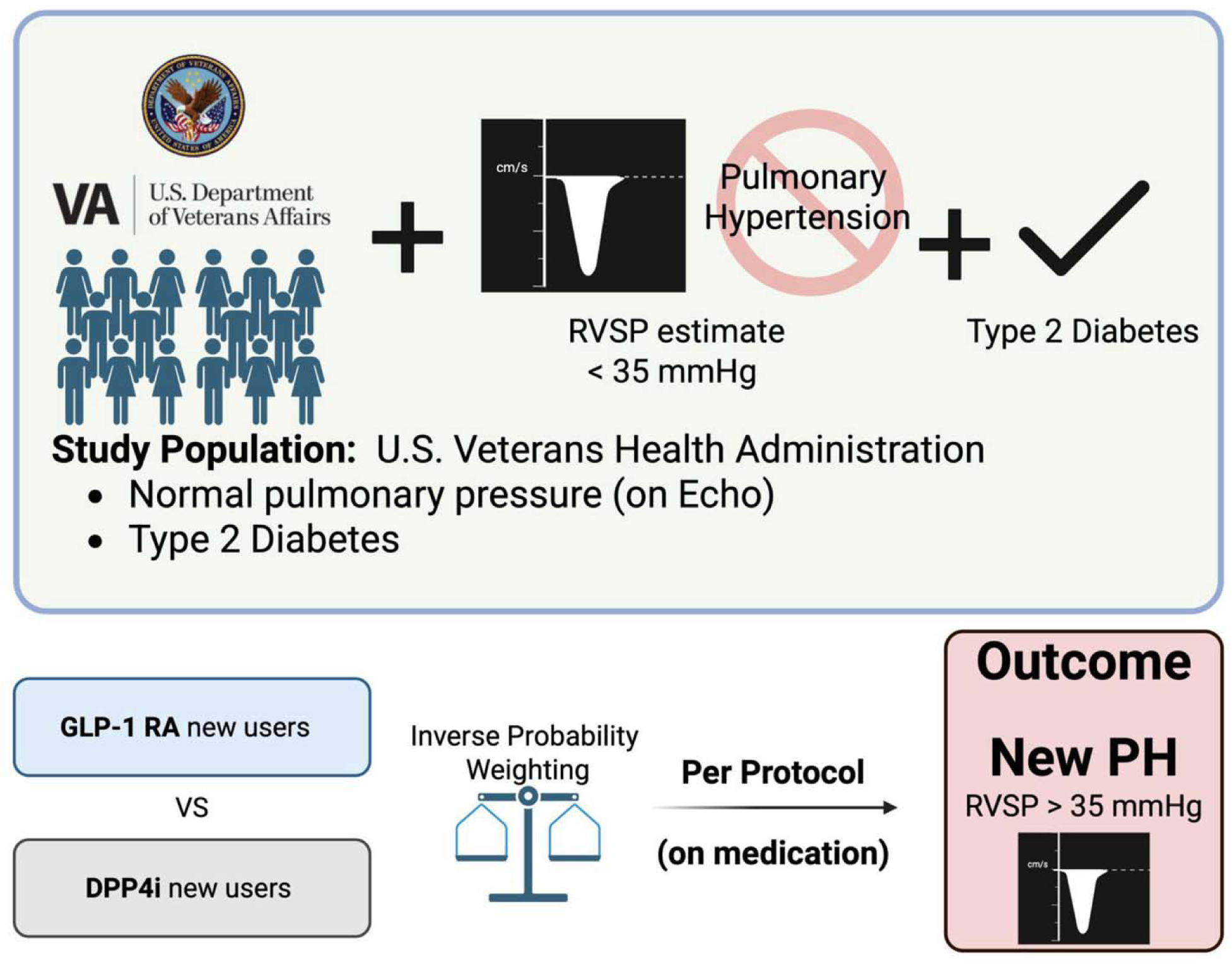
Study Design Overview Figure 1 displays the main features of the study design. The study population is derived from patients seeking care at the U.S. Veterans Health Administration with a diagnosis of type 2 diabetes and normal pulmonary pressure on echocardiography who are newly prescribed a Glucagon Like Peptide 1 Receptor Agonist (GLP-1 RA) or a Dipeptidyl Peptidase 4 inhibitor (DPP4i). GLP-1 RA new users were then compared to DPP4i using inverse probability weighting to adjust for characteristics that influence medication selection. We followed patients while on-medication in a pre-protocol fashion until death, cessation of study drug, or development of new pulmonary hypertension, defined by an elevated echocardiographic right ventricular systolic pressure estimate of 35 mmHg or greater.

We identified all patients over 18 years with type 2 diabetes defined by ICD-9/10 codes and an echo without pulmonary hypertension (PH). PH was defined by a right ventricular systolic pressure (RVSP) estimate of ≥35 mmHg. Patients who initiated a GLP-1 RA (albiglutide, dulaglutide, exenatide, lixisenatide, liraglutide, or semaglutide) or DPP-4i (alogliptin, linagliptin, saxagliptin, or sitagliptin) after an echo that demonstrated the absence of PH prior to medication initiation were eligible for inclusion. The baseline date of study entry was defined by the date of first dispensed prescription for GLP-1 RA or DPP-4i. We then excluded patients with an echo demonstrating PH at any point prior to baseline and patients without at least one additional RVSP measurement after baseline. Patients were also excluded if they received a prescription for GLP-1 RA or DPP4i before the echo without PH and within 1 year of the baseline prescription. The primary outcome was the development of new PH on echo, defined by an RVSP ≥35 mmHg.

RVSP was directly abstracted from echo reports with a high-degree of precision, found to be 95-100% on manual validation.^11^ When RVSP was missing, but a tricuspid regurgitant velocity was reported, we calculated RVSP using the simplified Bernoulli equation. We assumed a RAP of 5 mmHg (median value in the general population) when missing.^14^

### Statistical Analysis and Inverse Probability Weighting (IPW)

We used propensity score weighting to balance baseline covariates. Patients in the GLP-1 RA group were weighted so that the covariate distributions matched those of the DPP-4i group (average treatment effect among treated weighting). The propensity scores were computed using logistic regression of exposure (GLP-1 RA vs DPP-4i) by baseline covariates.^15^ The distribution of the covariates was assessed by standardized mean differences (SMDs) between the treatment groups before and after weighting, with a value of less than 0.10 indicating good balance.^16^ In the propensity score model, continuous predictors were included as restricted cubic splines with three knots. For the propensity score model only, missing data in continuous variables were handled by a single imputation to each variable’s mean value, a point of low leverage in the model, and the inclusion of missingness indicator variables, which adjust the model’s intercept. For categorical variables, missing was added as a category. This approach balances both observable characteristics and missingness patterns between the treatment groups.^17,18^

Based on prior work and clinical knowledge, we selected the variables most likely to associate with the hazard of developing PH.^2^ These included: 1) Demographic characteristics including age and sex. 2) Vital signs including systolic blood pressure, diastolic blood pressure, and body mass index closest to baseline. 3) Comorbidities including heart failure, chronic obstructive pulmonary disease, atrial fibrillation, scleroderma, human immunodeficiency virus, and hepatitis C whose presence was determined by least two outpatient or one inpatient ICD-9/10 code as in prior studies (eTable 1).^2,19–21^ 4) Laboratory data including estimated glomerular filtration rate (eGFR, calculated from creatinine using the 2009 CKD-EPI equation^22^), hemoglobin concentration, and FIB-4 score (a composite of age, platelet count, aspartate aminotransferase and alanine aminotransferase used for prediction of liver fibrosis).^23,24^ 5) Medication use (active prescription) beginning 2 years prior up until 6 months after baseline, which was defined as present or absent for each medication class. Medication classes included loop diuretics (bumetanide, torsemide, and furosemide) and sodium-glucose cotransporter-2 inhibitors (bexagliflozin, canagliflozin, dapagliflozin, empagliflozin, ertugliflozin). 6) Echo related variables including left ventricular ejection fraction and time from echo to baseline. 7) Diabetes severity was addressed by including baseline insulin use, baseline hemoglobin A1c and time from diabetes diagnosis to baseline. 8) Smoking status, classified as current, former, or never. Comorbidities were considered present if noted any time before baseline up to 1 month after. All remaining variables were abstracted closest to baseline, beginning from 2 years prior up until 6 months after.

We generated Kaplan Meier curves for each IPW medication group. We assessed whether GLP-1 RAs were associated with hazard of developing PH compared to DPP-4i using Cox regression on the IPW cohort. We used robust standard errors adjusted for the IPW using the survey package with clustering by patient. In these analyses we treat death as a censoring criterion, not a competing risk. Thus, all hazard ratios in this paper refer to cause-specific hazard ratios, which better reflect the etiological association between GLP-1 RA use and PH. Estimators such as the subdistribution hazard ratio that account for competing risks are better for estimating measures of absolute increase. As GLP-1 RA are known to associate with lower all-cause mortality relative to DPP4i, the cause-specific hazard ratios provide the cleaner interpretation.^25–27^

### Primary, Sensitivity, and Subgroup Analyses

The primary analysis was conducted using a persistent exposure approach in which patients were followed from their initial medication exposure until they stopped receiving that medication (including 1-month after the end of the final refill), transitioned to the alternate medication, developed PH, or reached the end of the study period.

We conducted the following sensitivity analyses to ensure the robustness and consistency of our findings including: 1) In addition to IPW, we adjusted for covariates. For these models missing values were imputed using multiple imputation by chained equations with twenty separate imputed datasets based on predictive mean matching using the “mice” R package.^28^ Cox proportional hazards models were fitted in each imputed dataset and combined to obtain pooled hazard ratios and standard errors according to Rubin’s rules;^29^ 2) Modeling an intention-to-treat approach where we assumed the treatment status defined at baseline continued until the end of study (April 30, 2022). For this model we did not censor patients for cessation of medication or crossover; 3) Allowing for patients to contribute multiple episodes with a washout period of 1 year between episodes; 4) Censoring at the time of follow up echocardiogram; 5) Censoring at 4.5 years of follow up to ensure the robustness of our model at shorter and longer durations of follow-up. This threshold was selected due to a notable decline in the number of participants remaining under observation beyond this time point. 6) Completing primary analysis among only those patients without PH on an echo within 2 years of drug initiation. As the population for this analysis differed from the primary analysis IPW was completed again, de novo.

We conducted exploratory pre-specified subgroup analyses by age (>65 years vs ≤65 years), sex, body mass index (>30 kg/m^2^ vs ≤30 kg/m^2^), chronic obstructive pulmonary disease status, diabetes severity (HbA1c ≥8 vs <8), eGFR (≤60 vs >60), heart failure status with further stratification by left ventricular ejection fraction (≥50% vs <50%), and medication start date, comparing before vs after 2014, the year in which liraglutide was first approved for the indication of weight loss.

All analysis were conducted using R software (version 4.3.1; www.r-project.org). A two-sided p-value of less than 0.05 was considered statistically significant.

## Results

There were 30,671 patients with a diagnosis of diabetes who were newly prescribed a GLP-1 RA or DPP-4i on or after January 2007 and had an echo without PH (RVSP <35mmHg) before the medication start date. From this population, we identified 7,384 new DPP-4i starts and 4,109 new GLP-1 RA starts (eFigure 1). The distribution of prescriptions for individual GLP-1 RA and DPP-4i medications is provided in eTable 2, with histograms for the number of GLP-1 and DPP-4i prescriptions per year displayed in eFigure 2.

**Figure 2:**
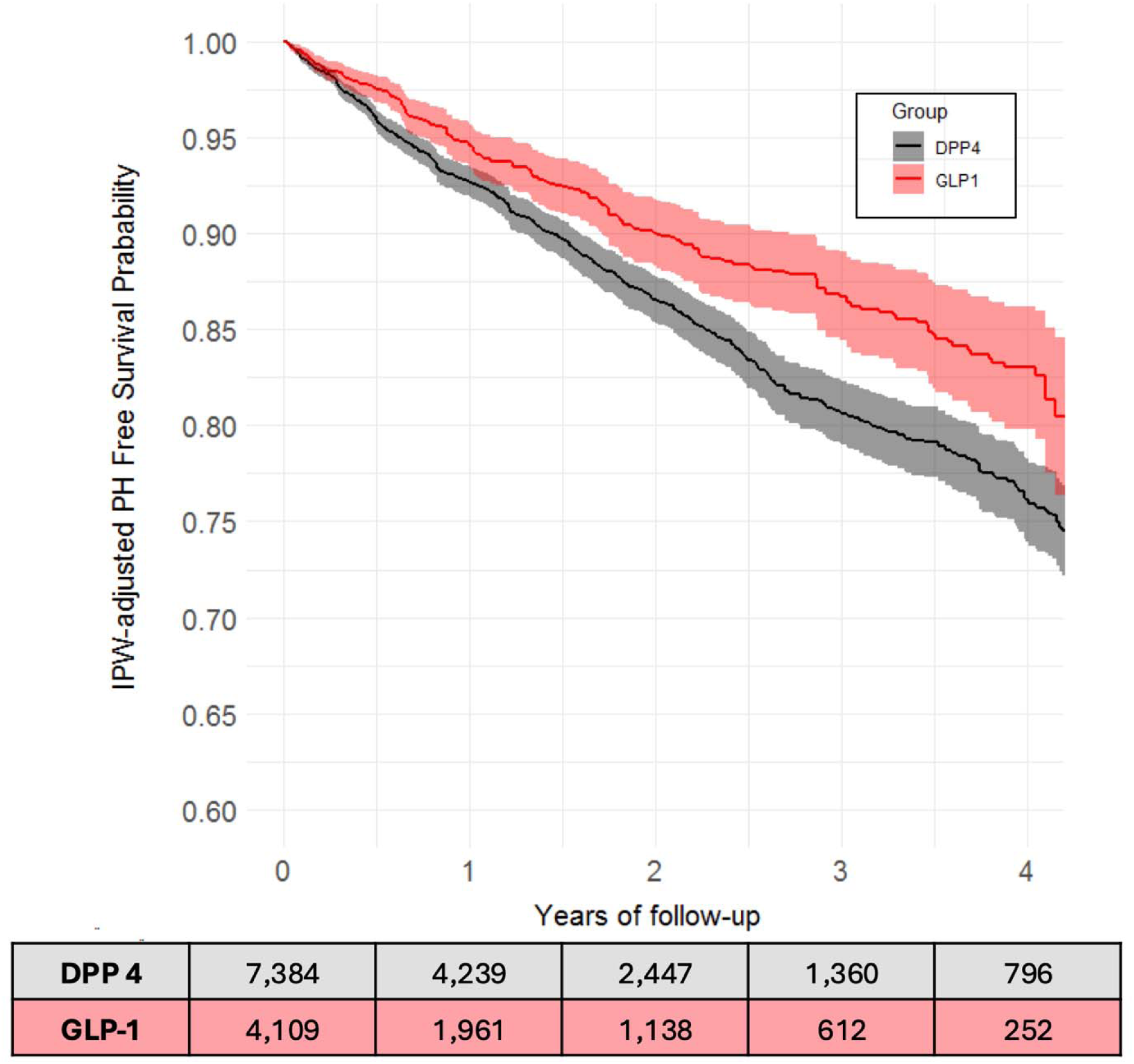
Figure 2 displays the Kaplan Meier curves for pulmonary hypertension free survival probability over time in the inverse probability weighted DPP4i (black) and GLP1-RA (red) cohorts. The at-risk table displays events the true number at risk from the unadjusted cohort.

GLP-1 RA new users tended to be younger (median age 68.8 years vs 70.0 years) with a higher proportion of females (5.4% vs 4.4%). GLP-1 RA new users also tended to have a higher median BMI (34.0 vs 31.2), a lower median eGFR (67 vs 70) and a higher prevalence of comorbidities that associate with incident PH including heart failure (45.5% vs 33.4%) chronic obstructive pulmonary disease (43.2% vs 41.1%) and atrial fibrillation (28.2% vs 25.5%), among others. The initial RVSP was similar between groups (27.5 vs 28.0 mmHg). The baseline characteristics of each group are displayed in Table 1. After IPW the two groups were well balanced across all covariates, as evidenced by absolute standardized mean differences below the cutoff of 0.1.

**Table 1:** Characteristics of the Patients at Baseline Before and After IPW Adjustment.

|  | Unadjusted |  |  | IPW Adjusted |  |  |
| --- | --- | --- | --- | --- | --- | --- |
|  | GLP-1 RA<br>(n = 4,109) | DPP-4i<br>(n = 7,384) | SMD | GLP-1 RA<br>(n = 4,109) | DPP-4i<br>(n = 4,271) | SMD |
| **Demographics** |  |  |  |  |  |  |
| Age (years) | 68.8 (63.1-72.7) | 70.0 (64.8-74.4) | 0.235 | 68.8 (63.1-72.7) | 69.0 (62.8-72.8) | 0.005 |
| Male Sex | 3886 (94.6%) | 7059 (95.6%) | 0.047 | 3886 (94.6%) | 3049 (94.6%) | <0.001 |
| Race |  |  | 0.085 |  |  | 0.101 |
| Black | 691 (16.8%) | 1349 (18.3%) |  | 691 (16.8%) | 823 (19.3%) |  |
| White | 3048 (74.2%) | 5216 (70.6%) |  | 3048 (74.2%) | 2981 (69.8%) |  |
| Other | 283 (7.1%) | 649 (8.8%) |  | 283 (7.1%) | 357 (8.4%) |  |
| Missing | 77 (1.9%) | 170 (2.3%) |  | 77 (1.9%) | 110 (2.6%) |  |
| **Vital Signs** |  |  |  |  |  |  |
| BMI (kg/m<sup>2</sup>) | 34.0 (30.0-38.4) | 31.3 (27.8-35.8) | 0.490 | 34.0 (30.0-38.4) | 34.7 (30.8-39.4) | 0.032 |
| BMI Missing | 15 (0.4%) | 21 (0.3%) | 0.014 | 15 (0.4%) | 18 (0.4%) | 0.009 |
| SBP (mmHg) | 128 (118-138) | 130 (119-139) | 0.101 | 128 (118-138) | 128 (117-138) | 0.016 |
| SBP Missing | 19 (0.5%) | 32 (0.4%) | 0.004 | 19 (0.5%) | 23 (0.5%) | 0.012 |
| DBP (mmHg) | 73.0 (66.0-80.0) | 73.0 (66.0-80.0) | 0.031 | 73.0 (66.0-80.0) | 73 (67-80) | 0.002 |
| DBP Missing | 19 (0.5%) | 32 (0.4%) | 0.004 | 19 (0.5%) | 23 (0.5%) | 0.012 |
| **Echo Variables** |  |  |  |  |  |  |
| LVEF (%) | 56.0 (46.0-62.5) | 57.5 (49.0-62.5) | 0.074 | 56.0 (46.0-62.5) | 55.0 (47.0-61.0) | 0.041 |
| LVEF Missing | 543 (13.2%) | 1344 (18.2%) | 0.137 | 543 (13.2%) | 568 (13.3%) | 0.002 |
| RVSP (mmHg) | 27.5 (22.5-31.7) | 28.0 (23.3-32.0) | 0.081 | 27.5 (22.5-31.7) | 27.5 (22.8-32.0) | 0.021 |
| **Comorbidities** |  |  |  |  |  |  |
| COPD | 1776 (43.2%) | 3038 (41.1%) | 0.042 | 1776 (43.2%) | 1811 (42.5%) | 0.016 |
| HF | 1864 (45.4%) | 2468 (33.4%) | 0.246 | 1864 (45.4%) | 1961 (45.9%) | 0.011 |
| AF | 1160 (28.2%) | 1882 (25.5%) | 0.062 | 1160 (28.2%) | 1235 (28.9%) | 0.015 |
| Scleroderma | 145 (3.5%) | 262 (3.5%) | 0.001 | 145 (3.5%) | 138 (3.2%) | 0.016 |
| HIV | 21 (0.5%) | 47 (0.6%) | 0.017 | 21 (0.5%) | 25 (0.6%) | 0.009 |
| Hepatitis C |  |  | 0.130 |  |  | 0.009 |
| Positive | 151 (3.7%) | 369 (5.0%) |  | 151 (3.7%) | 161 (3.8%) |  |
| Negative | 3747 (91.2%) | 6438 (87.2%) |  | 3747 (91.2%) | 3898 (91.3%) |  |
| Unknown | 211 (5.1%) | 577 (7.8%) |  | 211 (5.1%) | 212 (5.0%) |  |
| Smoking |  |  | 0.167 |  |  | 0.008 |
| Never | 1406 (34.2%) | 2372 (32.1%) |  | 1406 (34.2%) | 1472 (34.5%) |  |
| Current | 586 (14.3%) | 1230 (16.7%) |  | 586 (14.3%) | 607 (14.2%) |  |
| Former | 1783 (43.4%) | 2867 (38.8%) |  | 1783 (43.4%) | 1853 (43.4%) |  |
| Missing | 334 (8.1%) | 915 (12.4%) |  | 334 (8.1%) | 339 (7.9%) |  |
| **Medication Use** |  |  |  |  |  |  |
| Insulin | 3240 (78.9%) | 2779 (37.6%) | 0.920 | 3240 (78.9%) | 3421 (80.1%) | 0.031 |
| Loop Diuretic | 1907 (46.4%) | 2473 (33.5%) | 0.266 | 1907 (46.4%) | 2014 (47.2%) | 0.015 |
| SGLT2i | 910 (22.1%) | 652 (8.8%) | 0.374 | 910 (22.1%) | 1027 (24.1%) | 0.045 |
| **Labs** |  |  |  |  |  |  |
| Hg (mg/dL) | 13.6 (12.4-14.6) | 13.6 (12.5-14.7) | 0.053 | 13.6 (12.4-14.6) | 13.7 (12.5-14.8) | 0.024 |
| Hg Missing | 59 (1.4%) | 72 (1.0%) | 0.042 | 59 (1.4%) | 80 (1.9%) | 0.035 |
| Fib-4 | 1.5 (1.1-2.0) | 1.4 (1.0-1.9) | 0.121 | 1.5 (1.1-2.0) | 1.4 (1.0-1.9) | 0.015 |
| Fib-4 Missing | 86 (2.1%) | 157 (2.1%) | 0.002 | 86 (2.1%) | 108 (2.5%) | 0.030 |
| HbA1c (mmol/mol) | 7.7 (6.9-8.5) | 7.7 (6.9-8.7) | 0.048 | 7.7 (6.9-8.5) | 7.7 (6.9-8.7) | 0.009 |
| HbA1c Missing | 34 (0.8%) | 24 (0.3%) | 0.066 | 34 (0.8%) | 54 (1.3%) | 0.044 |
| eGFR (mL/min/1.73m<sup>2</sup>) | 70 (53-87) | 67 (50-85) | 0.091 | 70 (53-87) | 66 (50-84) | 0.020 |
| eGFR Missing | 16 (0.4%) | 16 (0.2%) | 0.031 | 16 (0.4%) | 16 (0.4%) | 0.027 |

GLP-1 RA = Glucagon-like peptide 1 receptor agonists; DPP4i = Dipeptidyl Peptidase 4 inhibitor; IPW = Inverse Probability Weight; RVSP = Right Ventricular Systolic Pressure; BMI = Body Mass Index; SBP = Systolic Blood Pressure; DBP = Diastolic Blood Pressure; LVEF = Left Ventricular Ejection Fraction; RVSP = Right Ventricular Systolic Pressure; COPD = Chronic Obstructive Pulmonary Disease; HF = Heart Failure; AF = Atrial Fibrillation; HIV = Human Immunodeficiency Virus; SGLT2i = Sodium-Glucose Cotransporter-2 inhibitor; Hg = Hemoglobin; HbA1c = Hemoglobin A1c (glycated hemoglobin); eGFR = Estimated Glomerular Filtration Rate.

Over 6,053 person-years of follow up, there were 332 cases of PH among new users of GLP-1 RAs, compared to 914 cases over 13,113 person-years of follow up in new users of DPP-4i new users. The unadjusted incidence rate of PH in the GLP-1 RA group was 54.8 cases per 1000 person-years compared to 69.7 cases per 1000 person-years in the DPP-4i group. The median time to follow-up RVSP measurement was 1.1 years (0.5–2.4 years) among patients who did not develop PH and 1.1 years (0.4–2.2 years) among patients who developed PH. The median change in right ventricular systolic pressure (RVSP) from baseline RVSP measurement was 0 mmHg (-1 to 9 mmHg) in GLP-1 RA users, and 0 mmHg (-1 to 8 mmHg) in DPP-4i users. Weight change from baseline was -5.5lbs per year (-15.3 to 0.4 lbs) among GLP-1 RA users and -2.3 lbs (-8.3 to 2.6lbs) among DPP-4i users. The rate of PH in the IPW-adjusted cohorts was higher in DPP-4i users compared to GLP-1 RA users, as displayed in Figure 2.

On Cox regression modeling of the IPW cohort, the use of GLP-1 RA was associated with a 28% lower risk of PH compared with the use of DPP-4is (Hazard Ratio 0.72; 95% CI, 0.61–0.84). The finding of decreased risk of PH associated with GLP-1 RA usage was consistent across sensitivity analyses, including when employing intention-to-treat modeling (Hazard Ratio 0.85; 95% CI, 0.76–0.95) or censoring at the time of follow up echo (Hazard Ratio 0.69; 95% CI, 0.59-0.81). The results of the primary and sensitivity analyses are displayed in Table 2.

**Table 2.**
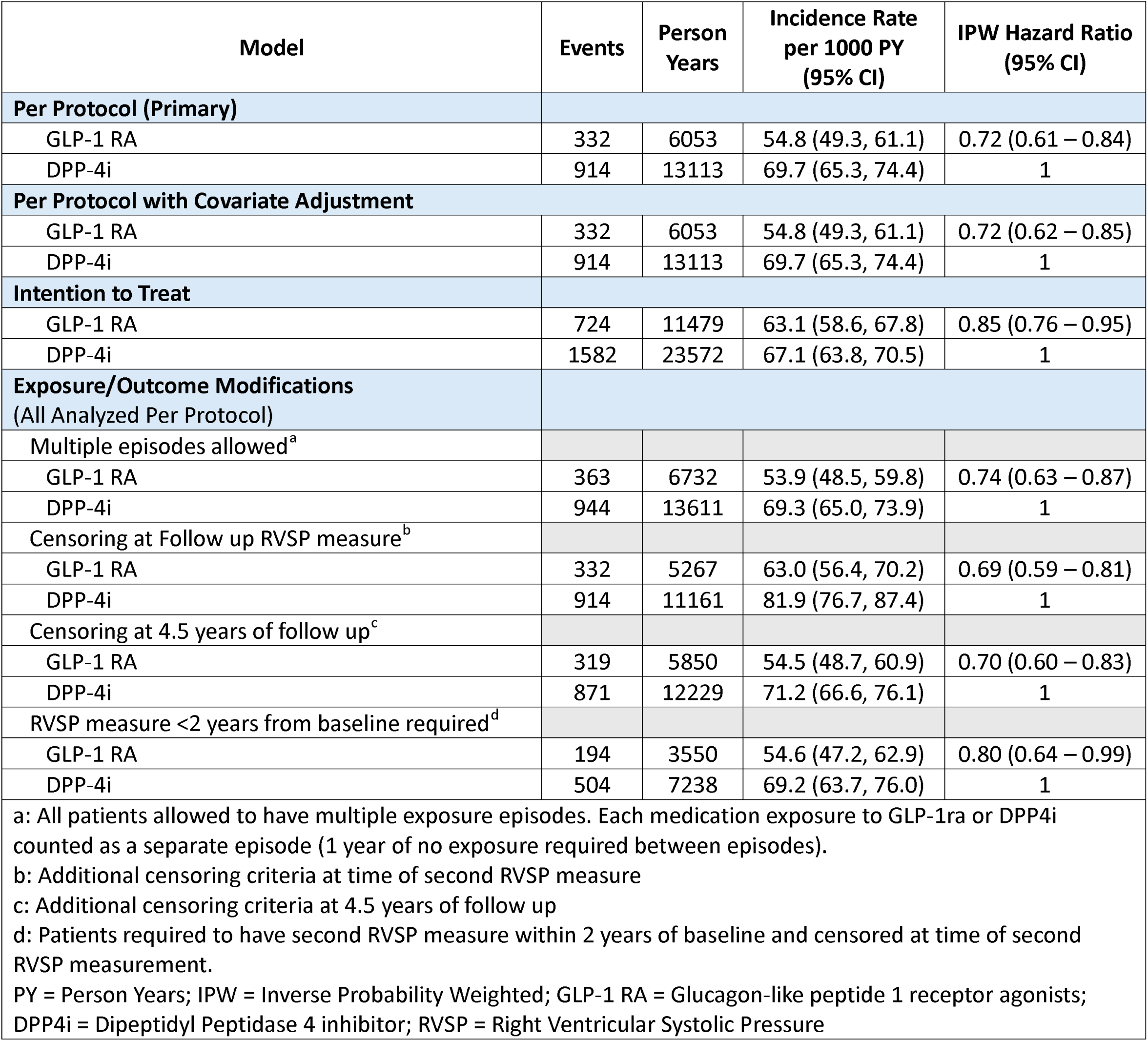
Primary and Sensitivity Model Results.

| Model | Events | Person Years | Incidence Rate per 1000 PY (95% CI) | IPW Hazard Ratio (95% CI) |
| --- | --- | --- | --- | --- |
| **Per Protocol (Primary)** |  |  |  |  |
| GLP-1 RA | 332 | 6053 | 54.8 (49.3, 61.1) | 0.72 (0.61 – 0.84) |
| DPP-4i | 914 | 13113 | 69.7 (65.3, 74.4) | 1 |
| **Per Protocol with Covariate Adjustment** |  |  |  |  |
| GLP-1 RA | 332 | 6053 | 54.8 (49.3, 61.1) | 0.72 (0.62 – 0.85) |
| DPP-4i | 914 | 13113 | 69.7 (65.3, 74.4) | 1 |
| **Intention to Treat** |  |  |  |  |
| GLP-1 RA | 724 | 11479 | 63.1 (58.6, 67.8) | 0.85 (0.76 – 0.95) |
| DPP-4i | 1582 | 23572 | 67.1 (63.8, 70.5) | 1 |
| **Exposure/Outcome Modifications (All Analyzed Per Protocol)** |  |  |  |  |
| Multiple episodes allowed<sup>a</sup> |  |  |  |  |
| GLP-1 RA | 363 | 6732 | 53.9 (48.5, 59.8) | 0.74 (0.63 – 0.87) |
| DPP-4i | 944 | 13611 | 69.3 (65.0, 73.9) | 1 |
| Censoring at Follow up RVSP measure<sup>b</sup> |  |  |  |  |
| GLP-1 RA | 332 | 5267 | 63.0 (56.4, 70.2) | 0.69 (0.59 – 0.81) |
| DPP-4i | 914 | 11161 | 81.9 (76.7, 87.4) | 1 |
| Censoring at 4.5 years of follow up<sup>c</sup> |  |  |  |  |
| GLP-1 RA | 319 | 5850 | 54.5 (48.7, 60.9) | 0.70 (0.60 – 0.83) |
| DPP-4i | 871 | 12229 | 71.2 (66.6, 76.1) | 1 |
| RVSP measure <2 years from baseline required<sup>d</sup> |  |  |  |  |
| GLP-1 RA | 194 | 3550 | 54.6 (47.2, 62.9) | 0.80 (0.64 – 0.99) |
| DPP-4i | 504 | 7238 | 69.2 (63.7, 76.0) | 1 |

a: All patients allowed to have multiple exposure episodes. Each medication exposure to GLP-1ra or DPP4i counted as a separate episode (1 year of no exposure required between episodes).
b: Additional censoring criteria at time of second RVSP measure
c: Additional censoring criteria at 4.5 years of follow up
d: Patients required to have second RVSP measure within 2 years of baseline and censored at time of second RVSP measurement.
PY = Person Years; IPW = Inverse Probability Weighted; GLP-1 RA = Glucagon-like peptide 1 receptor agonists; DPP4i = Dipeptidyl Peptidase 4 inhibitor; RVSP = Right Ventricular Systolic Pressure

In exploratory subgroup analysis, the effect of GLP-1 RA compared to DPP-4i appeared generally consistent when stratified by BMI, heart failure status (further subdivided into reduced and preserved ejection fraction based on a left ventricular ejection fraction cutoff of 40%), chronic obstructive pulmonary disease status, diabetes severity, eGFR, and medication start date (Figure 3). It was less clear whether the effect of GLP-1 RA was consistent across age and sex subgroups. Notably, there was a smaller number of patients under 65 and female patients compared to those greater than 65 and male patients, respectively, which resulted in wider confidence intervals.

**Figure 3.**
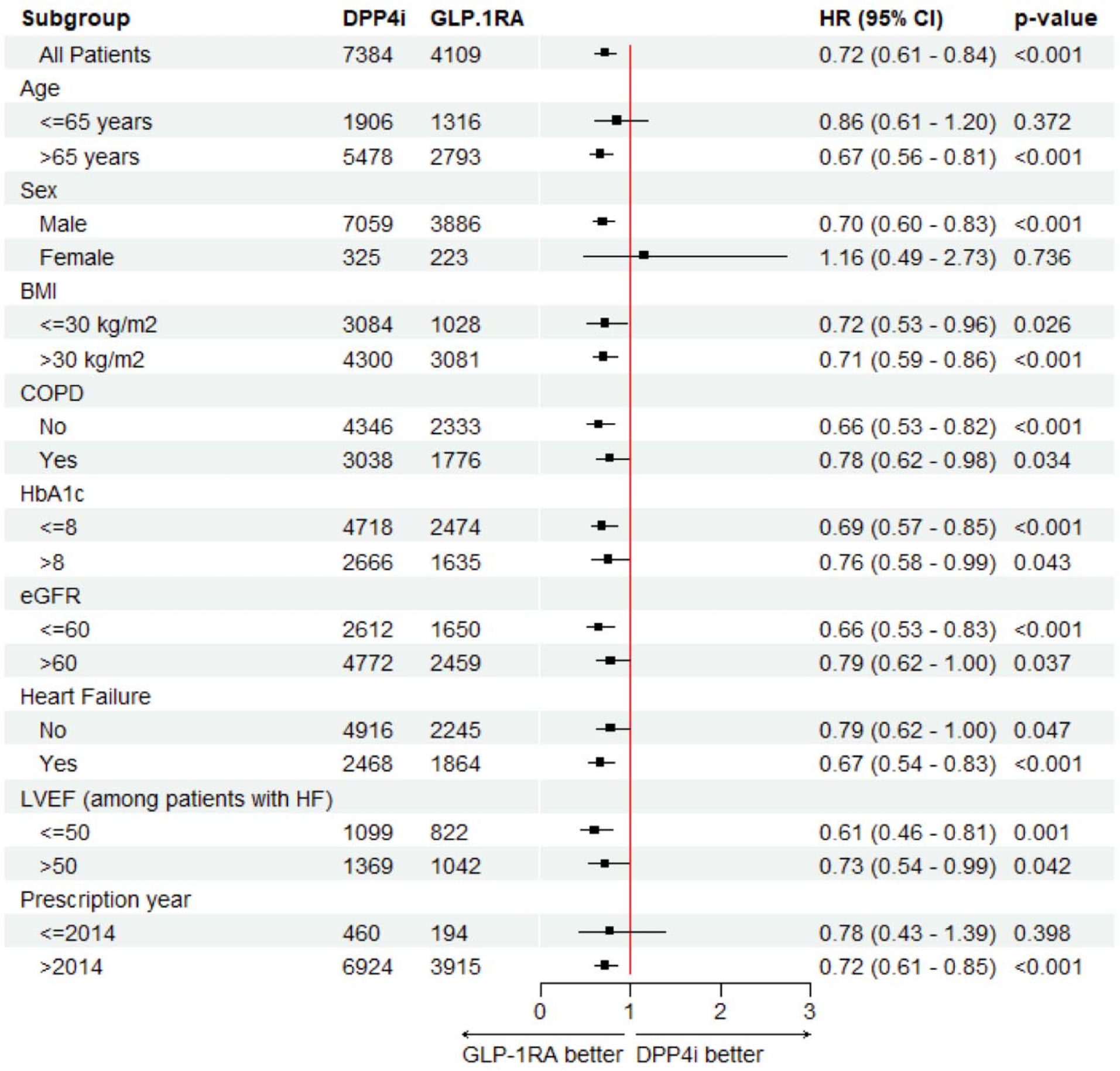
Forest Plot by Subgroups Figure 3 displays the subgroup analysis of the effect of GLP-1 RA exposure compared to DPP4i exposure on hazard of developing PH. BMI = Body mass index; COPD = Chronic obstructive pulmonary disease; HbA1c = Hemoglobin A1c; eGFR = estimated glomerular filtration rate; LVEF = Left ventricular ejection fraction; HF = Heart Failure

## Discussion

In this observational study using a clinical trial emulation design conducted in a large, diverse national cohort, we found that exposure to GLP-1 RAs was associated with a reduced hazard of developing PH, as defined by RVSP on echo. While GLP-1 RAs reduce the risk of major adverse cardiovascular events and improve metabolic health in patients with diabetes, our findings add important support to their potential benefit in preventing the development of elevated pulmonary pressures.^30^ This is particularly significant given the lack of existing interventions that prevent PH, especially among individuals with HF or COPD – two conditions that cause most PH cases at the population level.^2^

Compared to one prior study that found that GLP-1 RA use associated with reduced hazard of incident PH as an ancillary finding, the current study did not rely solely upon ICD-codes to determine PH status, was better equipped to adequately address confounding related to PH as an outcome, and carefully curated GLP-1RA medication data.^31^ Our study used echo data to define PH, offering greater diagnostic specificity and accuracy^2,32^ and employed a rigorous design to address potential confounding with multiple sensitivity analyses. Although we did not directly demonstrate downstream clinical benefit of GLP-1 RA use beyond prevention of PH in this study, it is plausible that reducing the incidence of PH would lead to improved clinical outcomes, including lower rates of heart failure hospitalization and reduced mortality. This is based on prior evidence that even mildly elevated pulmonary pressures on echocardiography are strongly associated with adverse outcomes.^3,4^

Multiple mechanisms may underlie the reduction in PH risk associated with GLP-1 RA exposure. In our population of predominantly older men with obesity and a high prevalence of heart failure, this association may be driven by improvements in left heart failure, consistent with evidence that GLP-1 RAs improve cardiovascular outcomes in patients with heart failure with preserved ejection fraction and obesity.^33^ GLP-1 RAs also exert salutary effects on blood pressure, lipid profile, hemoglobin A1c, weight, and renal function – each of which is independently associated with PH development.^2,34–37^ Amelioration of these conditions may in turn prevent or improve left heart failure, thereby reducing pulmonary venous congestion. Alternatively, reduction in cardiometabolic disease burden may secondarily stabilize pulmonary vascular remodeling. Moreover, GLP-1 RAs may also act directly on the pulmonary vasculature, independent of their effects on cardiometabolic comorbidities. Preclinical studies in rodent models of PH have demonstrated that GLP-1 RA exposure can reduce pulmonary pressures, decrease systemic inflammation and improve endothelial function, the latter mediated in part through increased nitric oxide synthase and reduced endothelin-1 expression.^5,8,35^

Although exploratory, our subgroup analyses suggest that the protective association between GLP-1 RA use and PH risk is consistent across a range of comorbid conditions and disease severity, including heart failure, COPD, degree of glycemic control, baseline presence of obesity, and severity of renal disease. This consistency suggests that GLP-1 RAs might exert pulmonary vascular benefits that are not fully attributable to improvements in these conditions. These findings also raise the possibility that GLP-1 RAs could reduce the risk of PH in individuals without diabetes or heart failure, warranting investigation in broader populations. Ultimately, further studies are needed to clarify the mechanisms through which GLP-1 RAs reduce the risk of developing PH.

### Limitations

As in any retrospective pharmacoepidemiologic study, our findings are subject to potential biases. We attempted to mitigate confounding by employing IPW and restricting the analysis to new users of GLP-1 RAs or DPP-4is in an active comparator design. Nevertheless, residual confounding cannot be fully excluded. Our primary analysis censored follow up at the end of exposure. This approach was selected to isolate the on-treatment effect but does carry risk of informative censoring. On intention-to-treat modeling we found an attenuation of effect, which may reflect the presence of such bias, but may also be attributable to post-exposure dilution of treatment effect. Our outcome of PH is defined by echo. This definition is not the gold standard and therefore may result in misclassification. However, we would not expect misclassification to differ between the exposure groups. The timing and frequency of follow up echos were determined by clinical discretion rather than standardized protocols, introducing potential detection bias. However, we adjusted for a comprehensive set of characteristics that likely influence the likelihood of follow-up imaging. In addition, on sensitivity analysis with censoring at the time of follow up echo there was no change in the observed association between GLP-1 RA exposure and PH risk. Follow up time in our primary analysis was relatively short, which may be influenced by our definition of exposure, which allowed only a 1-month gap in prescription before censoring. This definition may have captured frequent pauses or changes in GLP-1 RA or DPP-4i prescription that could signal a higher rate of medication side effects, which we were unable to capture. Nevertheless, we observed a robust benefit with a median follow up of just over 1 year, suggesting benefit manifests relatively early after exposure. Finally, our study was drawn from the VA and comprised primarily male patients with a wide confidence interval for the effect of GLP-1 RAs among women. As such, our findings warrant validation in more sex balanced cohorts.

## Conclusions

In this cohort study of patients with diabetes and evidence of normal pulmonary pressures, exposure to GLP-1 RAs associated with a reduced risk of developing echo-defined PH. Our findings suggest a potential role for GLP-1 RA treatment to prevent and manage PH. Future clinical trials are warranted to confirm this observation and define the mechanism of benefit.

## Supporting information

Supplemental Online Content

## Data Availability

Due to US Department of Veterans Affairs (VA) regulations and our ethics agreements, the analytic data sets used for this study are not permitted to leave the VA firewall without a Data Use Agreement. This limitation is consistent with other studies based on VA data. However, VA data are made freely available to researchers with an approved VA study protocol.

## Authors’ Contributions

JG, SK, SE, RG, MF, and EB contributed to study conception and design. CA performed data acquisition and SK performed data analysis. JG drafted the manuscript. All authors participated in data interpretation, critically reviewed the manuscript, and approved the final manuscript submitted for publication.

## Data Access, Responsibility, and Analysis

MF and EB had full access to all the data in the study and take responsibility for the integrity of the data and the accuracy of the data analysis.

## Patient and VA acknowledgement

This work uses data provided by patients and collected by the VA as part of their care and support.

## VA Disclaimer

The views and opinions expressed in this manuscript are those of the authors and do not necessarily represent those of the Department of Veterans Affairs or the United States government

## CMS data acknowledgement

Support for VA/CMS data provided by the Department of Veterans Affairs, VA Health Services Research and Development Service, VA Information Resource Center (Project Numbers SDR 02-237 and 98-004)

## References

1. Mocumbi A, Humbert M, Saxena A, Jing ZC, Sliwa K, Thienemann F, Archer SL, Stewart S. Pulmonary hypertension. Nat Rev Dis Primers. 2024;10:1. doi: 10.1038/s41572-023-00486-7

2. Garry JD, Kundu S, Annis J, Alcorn C, Eden S, Smith E, Greevy R, Maron BA, Freiberg M, Brittain EL. Incidence of Pulmonary Hypertension in the Echocardiography Referral Population. Ann Am Thorac Soc. 2024. doi: 10.1513/AnnalsATS.202407-716OC

3. Huston JH, Maron BA, French J, Huang S, Thayer T, Farber-Eger EH, Wells QS, Choudhary G, Hemnes AR, Brittain EL. Association of Mild Echocardiographic Pulmonary Hypertension With Mortality and Right Ventricular Function. JAMA Cardiol. 2019;4:1112–1121. doi: 10.1001/jamacardio.2019.3345

4. Maron BA, Hess E, Maddox TM, Opotowsky AR, Tedford RJ, Lahm T, Joynt KE, Kass DJ, Stephens T, Stanislawski MA, et al. Association of Borderline Pulmonary Hypertension With Mortality and Hospitalization in a Large Patient Cohort: Insights From the Veterans Affairs Clinical Assessment, Reporting, and Tracking Program. Circulation. 2016;133:1240–1248. doi: 10.1161/CIRCULATIONAHA.115.020207

5. King NE, Brittain E. Emerging therapies: The potential roles SGLT2 inhibitors, GLP1 agonists, and ARNI therapy for ARNI pulmonary hypertension. Pulm Circ. 2022;12:e12028. doi: 10.1002/pul2.12028

6. Gaspari T, Liu H, Welungoda I, Hu Y, Widdop RE, Knudsen LB, Simpson RW, Dear AE. A GLP-1 receptor agonist liraglutide inhibits endothelial cell dysfunction and vascular adhesion molecule expression in an ApoE-/- mouse model. Diab Vasc Dis Res. 2011;8:117–124. doi: 10.1177/1479164111404257

7. Honda J, Kimura T, Sakai S, Maruyama H, Tajiri K, Murakoshi N, Homma S, Miyauchi T, Aonuma K. The glucagon-like peptide-1 receptor agonist liraglutide improves hypoxia-induced pulmonary hypertension in mice partly via normalization of reduced ET(B) receptor expression. Physiol Res. 2018;67:S175–S184. doi: 10.33549/physiolres.933822

8. Wang J, Yu M, Xu J, Cheng Y, Li X, Wei G, Wang H, Kong H, Xie W. Glucagon-like peptide-1 (GLP-1) mediates the protective effects of dipeptidyl peptidase IV inhibition on pulmonary hypertension. J Biomed Sci. 2019;26:6. doi: 10.1186/s12929-019-0496-y

9. Wei R, Ma S, Wang C, Ke J, Yang J, Li W, Liu Y, Hou W, Feng X, Wang G, et al. Exenatide exerts direct protective effects on endothelial cells through the AMPK/Akt/eNOS pathway in a GLP-1 receptor-dependent manner. Am J Physiol Endocrinol Metab. 2016;310:E947–957. doi: 10.1152/ajpendo.00400.2015

10. Xu J, Wang J, He M, Han H, Xie W, Wang H, Kong H. Dipeptidyl peptidase IV (DPP-4) inhibition alleviates pulmonary arterial remodeling in experimental pulmonary hypertension. Lab Invest. 2018;98:1333–1346. doi: 10.1038/s41374-018-0080-1

11. Patterson OV, Freiberg MS, Skanderson M, J Fodeh S, Brandt CA, DuVall SL. Unlocking echocardiogram measurements for heart disease research through natural language processing. BMC Cardiovasc Disord. 2017;17:151. doi: 10.1186/s12872-017-0580-8

12. Lund JL, Richardson DB, Stürmer T. The active comparator, new user study design in pharmacoepidemiology: historical foundations and contemporary application. Curr Epidemiol Rep. 2015;2:221–228. doi: 10.1007/s40471-015-0053-5

13. Anderluh M, Kocic G, Tomovic K, Kocic H, Smelcerovic A. DPP-4 inhibition: А novel therapeutic approach to the treatment of pulmonary hypertension? Pharmacol Ther. 2019;201:1–7. doi: 10.1016/j.pharmthera.2019.05.007

14. Choudhary G, Jankowich M, Wu WC. Elevated pulmonary artery systolic pressure predicts heart failure admissions in African Americans: Jackson Heart Study. Circ Heart Fail. 2014;7:558–564. doi: 10.1161/CIRCHEARTFAILURE.114.001366

15. Austin PC. Differences in target estimands between different propensity score-based weights. Pharmacoepidemiol Drug Saf. 2023;32:1103–1112. doi: 10.1002/pds.5639

16. Austin PC. Balance diagnostics for comparing the distribution of baseline covariates between treatment groups in propensity-score matched samples. Stat Med. 2009;28:3083–3107. doi: 10.1002/sim.3697

17. Blake HA, Leyrat C, Mansfield KE, Seaman S, Tomlinson LA, Carpenter J, Williamson EJ. Propensity scores using missingness pattern information: a practical guide. Stat Med. 2020;39:1641–1657. doi: 10.1002/sim.8503

18. D’Agostino RB, Rubin DB. Estimating and Using Propensity Scores with Partially Missing Data. Journal of the American Statistical Association. 2000;95:749–759. doi: 10.2307/2669455

19. White JR, Chang CC, So-Armah KA, Stewart JC, Gupta SK, Butt AA, Gibert CL, Rimland D, Rodriguez-Barradas MC, Leaf DA, et al. Depression and human immunodeficiency virus infection are risk factors for incident heart failure among veterans: Veterans Aging Cohort Study. Circulation. 2015;132:1630–1638. doi: 10.1161/CIRCULATIONAHA.114.014443

20. Filipkowski AM, Kundu S, Eden SK, Alcorn CW, Justice AC, So-Armah KA, Tindle HA, Wells QS, Beckman JA, Freiberg MS, et al. Association of HIV Infection and Incident Abdominal Aortic Aneurysm Among 143 001 Veterans. Circulation. 2023;148:135–143. doi: 10.1161/CIRCULATIONAHA.122.063040

21. Freiberg MS, Chang CH, Skanderson M, Patterson OV, DuVall SL, Brandt CA, So-Armah KA, Vasan RS, Oursler KA, Gottdiener J, et al. Association Between HIV Infection and the Risk of Heart Failure With Reduced Ejection Fraction and Preserved Ejection Fraction in the Antiretroviral Therapy Era: Results From the Veterans Aging Cohort Study. JAMA Cardiol. 2017;2:536–546. doi: 10.1001/jamacardio.2017.0264

22. Levey AS, Stevens LA, Schmid CH, Zhang YL, Castro AF, 3rd, Feldman HI, Kusek JW, Eggers P, Van Lente F, Greene T, et al. A new equation to estimate glomerular filtration rate. Ann Intern Med. 2009;150:604–612. doi: 10.7326/0003-4819-150-9-200905050-00006

23. Sterling RK, Lissen E, Clumeck N, Sola R, Correa MC, Montaner J, S Sulkowski M, Torriani FJ, Dieterich DT, Thomas DL, et al. Development of a simple noninvasive index to predict significant fibrosis in patients with HIV/HCV coinfection. Hepatology. 2006;43:1317–1325. doi: 10.1002/hep.21178

24. Long MT, Noureddin M, Lim JK. AGA Clinical Practice Update: Diagnosis and Management of Nonalcoholic Fatty Liver Disease in Lean Individuals: Expert Review. Gastroenterology. 2022;163:764–774.e761. doi: 10.1053/j.gastro.2022.06.023

25. Austin PC, Lee DS, Fine JP. Introduction to the Analysis of Survival Data in the Presence of Competing Risks. Circulation. 2016;133:601–609. doi: 10.1161/CIRCULATIONAHA.115.017719

26. Evans M, Kuodi P, Akunna CJ, McCreedy N, Donsmark M, Ren H, Nnaji CA. Cardiovascular and renal outcomes of GLP-1 receptor agonists vs. DPP-4 inhibitors and basal insulin in type 2 diabetes mellitus: A systematic review and meta-analysis. Diab Vasc Dis Res. 2023;20:14791641231221740. doi: 10.1177/14791641231221740

27. Kristensen SL, Rorth R, Jhund PS, Docherty KF, Sattar N, Preiss D, Kober L, Petrie MC, McMurray JJV. Cardiovascular, mortality, and kidney outcomes with GLP-1 receptor agonists in patients with type 2 diabetes: a systematic review and meta-analysis of cardiovascular outcome trials. Lancet Diabetes Endocrinol. 2019;7:776–785. doi: 10.1016/S2213-8587(19)30249-9

28. van Buuren S, Groothuis-Oudshoorn K. mice: Multivariate Imputation by Chained Equations in R. Journal of Statistical Software. 2011;45:1–67. doi: 10.18637/jss.v045.i03

29. Little RJA, Rubin DB, ProQuest. Statistical analysis with missing data. Second edition. ed. Hoboken: Wiley; 2002.

30. Marx N, Husain M, Lehrke M, Verma S, Sattar N. GLP-1 Receptor Agonists for the Reduction of Atherosclerotic Cardiovascular Risk in Patients With Type 2 Diabetes. Circulation. 2022;146:1882–1894. doi: 10.1161/CIRCULATIONAHA.122.059595

31. Xie Y, Choi T, Al-Aly Z. Mapping the effectiveness and risks of GLP-1 receptor agonists. Nat Med. 2025. doi: 10.1038/s41591-024-03412-w

32. Maron BA, Choudhary G, Khan UA, Jankowich MD, McChesney H, Ferrazzani SJ, Gaddam S, Sharma S, Opotowsky AR, Bhatt DL, et al. Clinical profile and underdiagnosis of pulmonary hypertension in US veteran patients. Circ Heart Fail. 2013;6:906–912. doi: 10.1161/CIRCHEARTFAILURE.112.000091

33. Packer M, Zile MR, Kramer CM, Baum SJ, Litwin SE, Menon V, Ge J, Weerakkody GJ, Ou Y, Bunck MC, et al. Tirzepatide for Heart Failure with Preserved Ejection Fraction and Obesity. N Engl J Med. 2025;392:427–437. doi: 10.1056/NEJMoa2410027

34. Perkovic V, Tuttle KR, Rossing P, Mahaffey KW, Mann JFE, Bakris G, Baeres FMM, Idorn T, Bosch-Traberg H, Lausvig NL, et al. Effects of Semaglutide on Chronic Kidney Disease in Patients with Type 2 Diabetes. N Engl J Med. 2024;391:109–121. doi: 10.1056/NEJMoa2403347

35. Nauck MA, Meier JJ, Cavender MA, Abd El Aziz M, Drucker DJ. Cardiovascular Actions and Clinical Outcomes With Glucagon-Like Peptide-1 Receptor Agonists and Dipeptidyl Peptidase-4 Inhibitors. Circulation. 2017;136:849–870. doi: 10.1161/CIRCULATIONAHA.117.028136

36. Robbins IM, Newman JH, Johnson RF, Hemnes AR, Fremont RD, Piana RN, Zhao DX, Byrne DW. Association of the metabolic syndrome with pulmonary venous hypertension. Chest. 2009;136:31–36. doi: 10.1378/chest.08-2008

37. Morrison AM, Huang S, Annis JS, Garry JD, Hemnes AR, Freiberg MS, Brittain EL. Cardiometabolic Risk Factors Associated With Right Ventricular Function and Compensation in Patients Referred for Echocardiography. J Am Heart Assoc. 2023;12:e028936. doi: 10.1161/JAHA.122.028936

