## Supplemental Online Content for "Glucagon-like Peptide 1 Receptor Agonists and Risk of Pulmonary Hypertension"

**Title**: GLP-1 Receptor Agonists and Risk of Pulmonary Hypertension

**eTable 1.** Comorbidity Definitions

**eTable 2.** Breakdown of GLP-1 RA and DPP4i Agents

**eFigure 1.** Study flow diagram

**eFigure 2.** Distribution of GLP-1 RA and DPP4i prescription by year

| **eTable 1 –** Comorbidity Definitions | | |
| --- | --- | --- |
| **Comorbidity** | ICD 9^th^ Revision | ICD 10^th^ Revision |
| **Heart Failure** | 402.01, 402.11, 402.91, 404.01, 404.03, 404.11, 404.13, 404.91, 404.93, 428.0, 428.1, 428.21, 428.22, 428.23, 428.31, 428.32, 428.33, 428.41, 428.42, 428.43, 428.9, 428.20, 428.30, 428.40 | I11.0, I13.0, I13.2, I50.1, I50.20, I50.21, I50.22, I50.23, I50.30, I50.31, I50.32, I50.33, I50.40, I50.41, I50.42, I50.43, I50.810, I50.811, I50.812, I50.813, I50.814, I50.82, I50.83, I50.84, I50.89,  I50.9 |
| **Chronic Obstructive Pulmonary Disease** | 490, 491.0, 491.1, 491.2, 491.20, 491.21, 491.22, 491.8, 491.9, 496, 492.8, 492.0 | J40.X, J41.X, J42.X, J43.X, J44.X |
| **Atrial Fibrillation** | 427.31 | I48.0, I48.1, I48.2, I48.3, I48.4, I48.9 |
| **Human Immunodeficiency Virus** | 42.X | V08.X, B20.X, Z21.X |
| **Scleroderma** | 695.4, 710.0, 710.1, 710.2, 710.3, 710.4, 710.8, 710.9, 714.0 | J99.0, J99.1, M32.X, M33.X, M34.X, M35.X, M05.X |
| **Hepatitis C** | 70.44, 70.51, 70.54, 70.70,70.71 | B18.2 |
| NOTE: ‘X’ indicates all numbers after decimal (0-9). Diagnosis established by the presence of one inpatient ICD 9/10 code or two outpatient ICD 9/10 codes, at the time of the second outpatient ICD code. | | |

| **eTable 2 –** Breakdown of GLP-1 RA and DPP4i Agents | | | | |
| --- | --- | --- | --- | --- |
| **GLP-1 RA**  **(n = 4,109)** | | | **DPP4i**  **(n = 7,384)** | |
| **Medication**  **Name** | **Number (%)** | **Medication**  **Name** | | **Number (%)** |
| Albiglutide | 3 (0.1%) | Alogliptin | | 3099 (42.0%) |
| Dulaglutide | 847 (20.6%) | Linagliptin | | 400 (5.4%) |
| Exenatide | 259 (6.3%) | Saxagliptin | | 3322 (45.0%) |
| Lixisenatide | 2 (<0.1%) | Sitagliptin | | 563 (7.6%) |
| Liraglutide | 1881 (45.8%) |  | |  |
| Semaglutide | 1117 (27.2%) |  | |  |
| GLP-1 RA = Glucagon-like peptide 1 receptor agonist; DPP4i = Dipeptidyl Peptidase 4 inhibitor | | | | |

**eFigure 1:** Study Flow Diagram

eFigure 1 displays the patient identification process including key elements of inclusion criteria, exclusion rationale, and final cohort size.

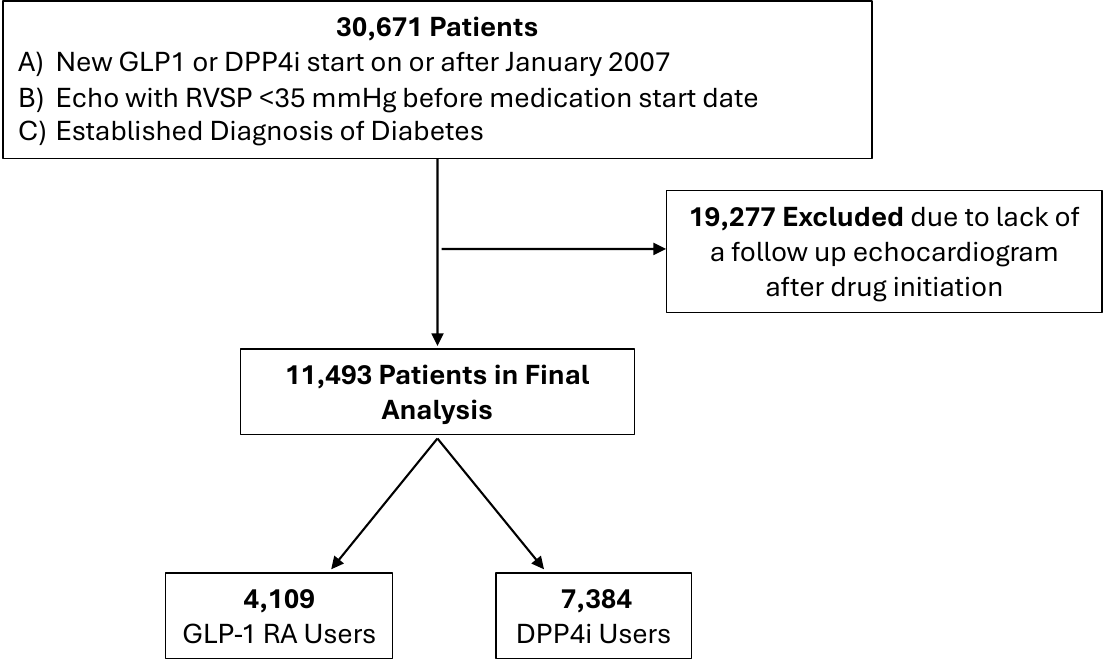

**eFigure 2:** Distribution of GLP-1 RA and DPP4i prescription by year

**eFigure 2** shows histograms displaying the baseline year of GLP-1 RA and DPP4i prescriptions for patients included in the study

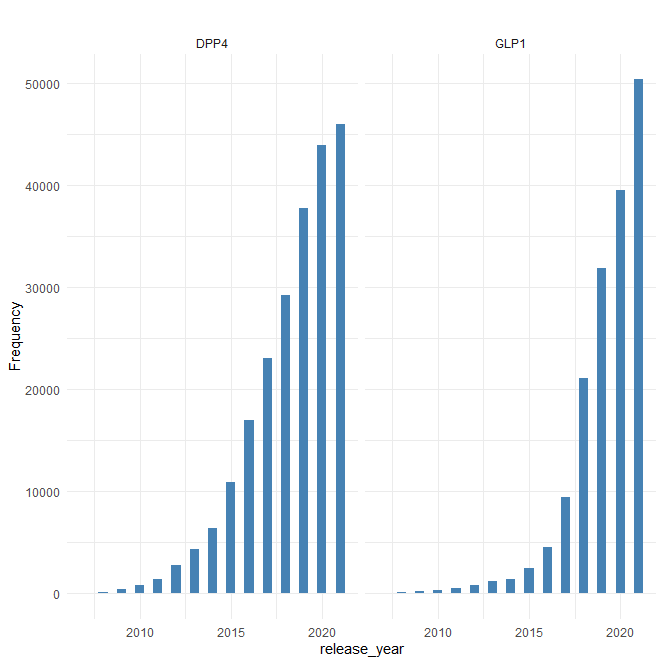

Prescription Year
